# Patient-level forecasting of geographic atrophy with a biologically grounded Gompertz model

**DOI:** 10.1101/2025.10.02.25337187

**Authors:** Aaron D. Beckwith, Stephen M. McNamara, Yoga Advaith Veturi, Niranjan Manoharan, Talisa E. de Carlo Forest, Scott Kinder, Benjamin Bearce, Ramya Gnanaraj, Anne Lynch, Praveer Singh, Giacomo Nebbia, Naresh Mandava, Jayashree Kalpathy–Cramer

## Abstract

We tested whether a Gompertz growth curve better describes and predicts geographic atrophy lesion enlargement than linear and effective-radius (square-root) models. We analyzed a retrospective, single-center cohort of 121 patients (181 eyes) with serial fundus autofluorescence imaging from October 2012 to April 2023, excluding eyes that had received prior geographic atrophy therapies or fewer than five gradable visits, creating a natural-history cohort. We fitted four candidate models (Gompertz, logistic, linear, and effective radius) within a hierarchical framework. We evaluated model accuracy using rolling out-of-sample forecasts, as assessed by continuous ranked probability scores. We assessed calibration by the prediction-interval width and coverage. The median follow-up was 5.8 years (IQR, 2.8 years), the mean age was 79.2 years (SD, 7.9 years), and 60% of the cohort were female. Gompertz achieved the lowest forecast error (0.45 mm^2^) versus logistic (0.48 mm^2^), linear (0.52 mm^2^), and effective radius (0.62 mm^2^), and received the highest pseudo-Bayesian model averaging weight (0.994). It yielded narrower 90% prediction intervals (2.41 mm^2^ vs. 3.99 mm^2^ for linear) and maintained these advantages at longer forecast horizons, where traditional models tended to overpredict. Differences were most pronounced during late (decelerating) growth. These findings demonstrate that Gompertz trajectories better capture lesion enlargement and modestly improve probabilistic forecasts compared with conventional approaches, supporting their use for patient counseling and for trial designs that account for natural growth deceleration.

## Introduction

Geographic atrophy (GA), a non-exudative late-stage of age-related macular degeneration (AMD), is a major cause of central vision loss, affecting millions of individuals among the 196 million worldwide living with AMD [1]. As GA progression often leads to irreversible blindness, predicting the rate and trajectory of lesion growth plays a pivotal role in guiding patient counseling, optimizing treatment timing, and designing effective randomized controlled trials (RCTs) [2–4]. Robust prognostic models of lesion growth are particularly critical at this juncture, as newly approved therapeutic agents such as complement inhibitors [5] emerge for this previously untreatable condition. Current GA progression models frequently fail to reflect the biological complexity of the disease, thereby limiting their clinical and research utility.

GA lesion assessment is mainly based on fundus examination, optical coherence tomography (OCT) [6] and fundus autofluorescence (FAF) [7] imaging, relying on a combination of metrics like lesion size and number with qualitative descriptors of shape, margin characteristics, and foveal involvement [8,9]. While somewhat clinically informative, this approach typically stops short of the dynamic, longitudinal modeling of nonlinear growth rates required for robust prognostication. By contrast, natural-history cohorts and late-phase RCTs employ a more rigorous standard, quantifying total GA lesion area in mm^2^ from standardized images as the primary anatomic efficacy endpoint [2–4, 10, 11]. Despite advancements in imaging technology and GA segmentation methods [12], existing quantitative models for GA progression typically rely on linear models of either the lesion area [13, 14] or the effective radius (square root of area) [15, 16]. Because these methods assume unbounded growth, they may bias results by ignoring biologically expected deceleration due to diminishing susceptible cell populations [17], altered tissue environments [18–20], and anatomical boundaries as lesions enlarge [21–23]. Although the necessity of bounded growth is well-established mathematically [23] and in other medical domains, such as oncology [24–28], these have not yet been adopted in the GA literature, representing a methodological gap. Consequently, recent evidence suggests the square-root transformation does not eliminate baseline lesion-size dependency from GA growth rates [29], indicating further shortcomings. These linear methods may not adequately account for patients’ natural disease phase, potentially confounding progression estimates when comparing eyes at different growth stages, a hypothesis that has not yet been empirically validated with clinical trial data.

This gap persists not due to technical barriers, but because linear approaches were historically sufficient for estimating average growth rates and comparing treatment effects with available datasets. However, as longer follow-up data becomes available and precision forecasting gains importance, particularly for detecting subtle drug effects, sigmoidal models offer clear advantages. The Gompertz function is particularly well-suited for GA progression because its asymmetric sigmoidal shape allows for early rapid expansion followed by gradual tapering consistent with progressive depletion of susceptible photoreceptor units, unlike symmetric logistic curves that assume sharp mid-course deceleration.

To address these limitations, we developed a framework integrating AI-driven segmentation of GA from FAF images with advanced mathematical growth models (Fig. 1). We compared established linear and effective-radius models against nonlinear sigmoidal logistic [30–32] and Gompertz [24–27, 33] models, which explicitly account for biologically expected growth deceleration and eventual saturation as lesions approach anatomical limits. We hypothesized that the Gompertz function, with its asymmetric shape reflecting how GA lesions typically grow rapidly initially then gradually decelerate as susceptible cell populations diminish, would outperform traditional methods, particularly for patients with sparse visit histories. As the first study we are aware of to validate the Gompertz model for GA progression using longitudinal patient data, we aimed to develop a robust prognostic tool to accurately forecast disease trajectories, thereby enhancing patient counseling, drug-response monitoring, and future RCT design.

**Fig 1.**
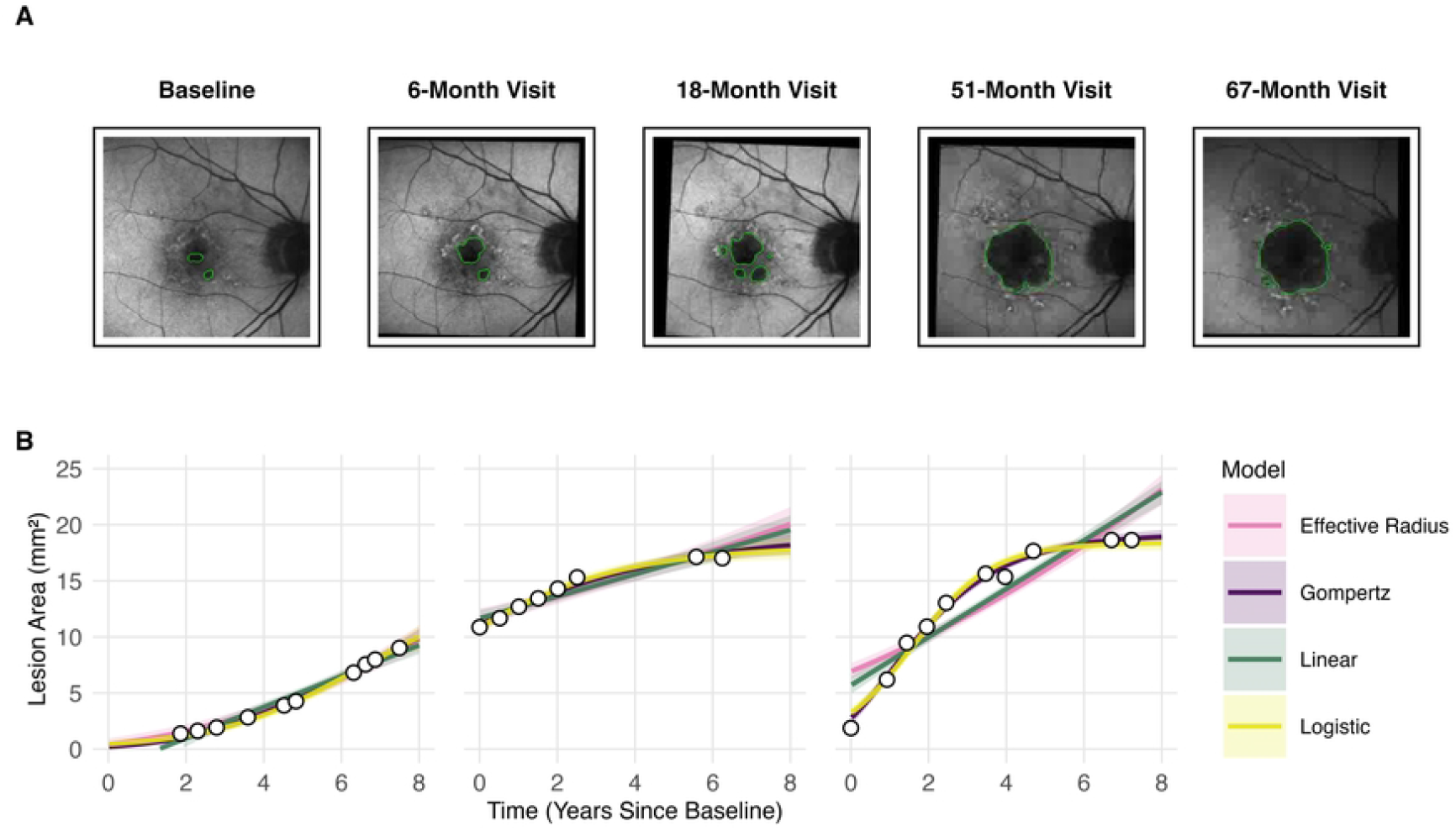
Segmentation, area extraction, and growth-curve analysis. (A) Retinal fundus-autofluorescence images were segmented with MedSAM [34] to delineate geographic atrophy and displayed with Eyeliner [35]. Lesion area (mm^2^) was quantified at each visit, generating longitudinal data. (B) Points represent observed lesion areas; colored lines depict posterior mean predictions with shaded 95% credible intervals. The Gompertz model (purple) captures the biologic deceleration phase, whereas the linear (teal) and effective-radius (pink) models remain unbounded, and the logistic model (yellow) imposes a symmetry that does not match the patient’s early rapid expansion.

## Methods

### Study design and population

We conducted a single-center, retrospective longitudinal cohort study at the Sue Anschutz Rodgers Eye Center using data from the Colorado Ophthalmology Research Information System (CORIS). From the electronic health records, we first identified 2,438 patients with a diagnosis of dry AMD and concurrent GA (ICD-10 H35.31x; ICD-9 362.51) who had FAF images acquired between October 2012 and April 2023. We excluded cases with concurrent neovascular AMD or prior GA-directed therapy to focus on natural history.

From this initial cohort, we included eyes if they had *≥*5 gradable FAF visits with at least three visits showing a lesion area *>* 2 mm^2^, ensuring a measurable growth signal above imaging noise and stabilizing asymptote estimates [15, 24–27, 33, 36–39]. After applying these criteria, 308 eyes (with 2,501 FAF images) remained. These images underwent quality control by three reviewers; images with motion artifacts, poor focus, or other quality issues affecting segmentation fidelity were excluded (20% of images) resulting in a final analytic cohort of 121 patients (181 eyes) with 1,433 images over a median follow-up of 5.8 years (IQR, 2.8 years). See detailed inclusion flowchart in S1 Fig. and an analysis of differences between included and excluded patients in S1 Appendix (higher female proportion among included; OR*≈* 2.1 with CrI excluding 1).

### Imaging acquisition and lesion segmentation

We acquired FAF images using standardized settings on Spectralis devices (Heidelberg Engineering GmbH; Heidelberg, Germany). The lesion area (mm^2^) was segmented using a novel deep-learning pipeline, based on the Medical Segment Anything Model (MedSAM) [34] fine-tuned on 304 clinician-annotated FAF images. On an independent test set (344 images), the area predictions from our algorithm showed near-perfect agreement with masked graders (Pearson r = 0.99).

### Growth model framework and statistical analysis

#### Model selection and description

Lesion area (mm^2^) for each eye was modeled over time since first clinic visit using four growth assumptions: Gompertz, logistic, linear, and effective radius. Gompertz and logistic models explicitly account for growth deceleration as lesions approach anatomical boundaries, while linear and square-root models assume unbounded growth (S2 Fig.). Full model specifications, equations, and Bayesian prior distributions are in the supplementary materials. Sensitivity analyses using von Bertalanffy and Mitscherlich curves yielded similar results (S8 Table).

#### Model fitting and estimation

Hierarchical Bayesian models captured intra- and interpatient variability, implemented using R (version 4.4.1, R Foundation for Statistical Computing) [40], brms (version 2.22.0) with cmdstanr (version 0.8.0) [41] interface. This approach jointly estimated key parameters, including growth rate, asymptotic lesion size, and scaling; additional computational details are included in S3 Appendix.

#### Metrics for model comparison

We evaluated performance using the continuous ranked probability score (CRPS) [42–46] to assess predictive accuracy across the full range of possible lesion areas, with lower values indicating better forecasting. We calculated posterior model probabilities using pseudo-Bayesian model averaging (pseudo-BMA^+^) [47], which combines predictive performance while penalizing overfitting to yield probabilities for each model being optimal for new data. We also calculated mean absolute error (MAE) as the average absolute difference between predicted medians and observed values [48] as a supplement.

#### Validation and prognostic accuracy (sequential forecasting)

To mimic real-world clinical scenarios with irregular follow-up, we performed a sequential forecasting validation. Starting from each patient’s fifth visit, we used all available historical data to predict lesion size at each subsequent visit, yielding 316 eye-specific forecasts. We assessed prognostic accuracy using CRPS and MAE from predictive medians and evaluated model calibration via prediction interval coverage and width.

To account for clustering and clinical heterogeneity in forecast conditions (some patients have variable follow-up times or number of clinical visits), we modeled the forecasting CRPS itself using a hierarchical regression: CRPS was treated as Gamma-distributed with a log link; the linear predictor included a fixed effect for growth model, a model-specific horizon term, the number of prior visits and its interaction with model, and random intercepts for patient and eye. This single framework produced all model-adjusted results reported, expected CRPS (with 95% CrIs) by model and horizon, marginal effects for horizon and data density, pairwise ΔCRPS, and the population-level posterior probability that each model yields the lowest expected CRPS for a new forecast.

Finally, to test if model accuracy varied by growth dynamics, we classified each forecasting window into distinct phases. Given that sigmoidal lesion expansion proceeds through distinct dynamic phases (initial acceleration, near-linear growth, then deceleration toward a plateau; see Fig. 2A), we anticipated that model accuracy would vary depending on where each observation window fell along this trajectory. Using the time of maximum growth rate and inflection points as mathematical landmarks, each window was categorized as acceleration phase (early rapid growth), linear phase (peak growth rate), deceleration phase (slowing growth), or mixed phase (spanning transitions). This classification enabled stratified evaluation of model performance across different stages of lesion development.

**Fig 2.**
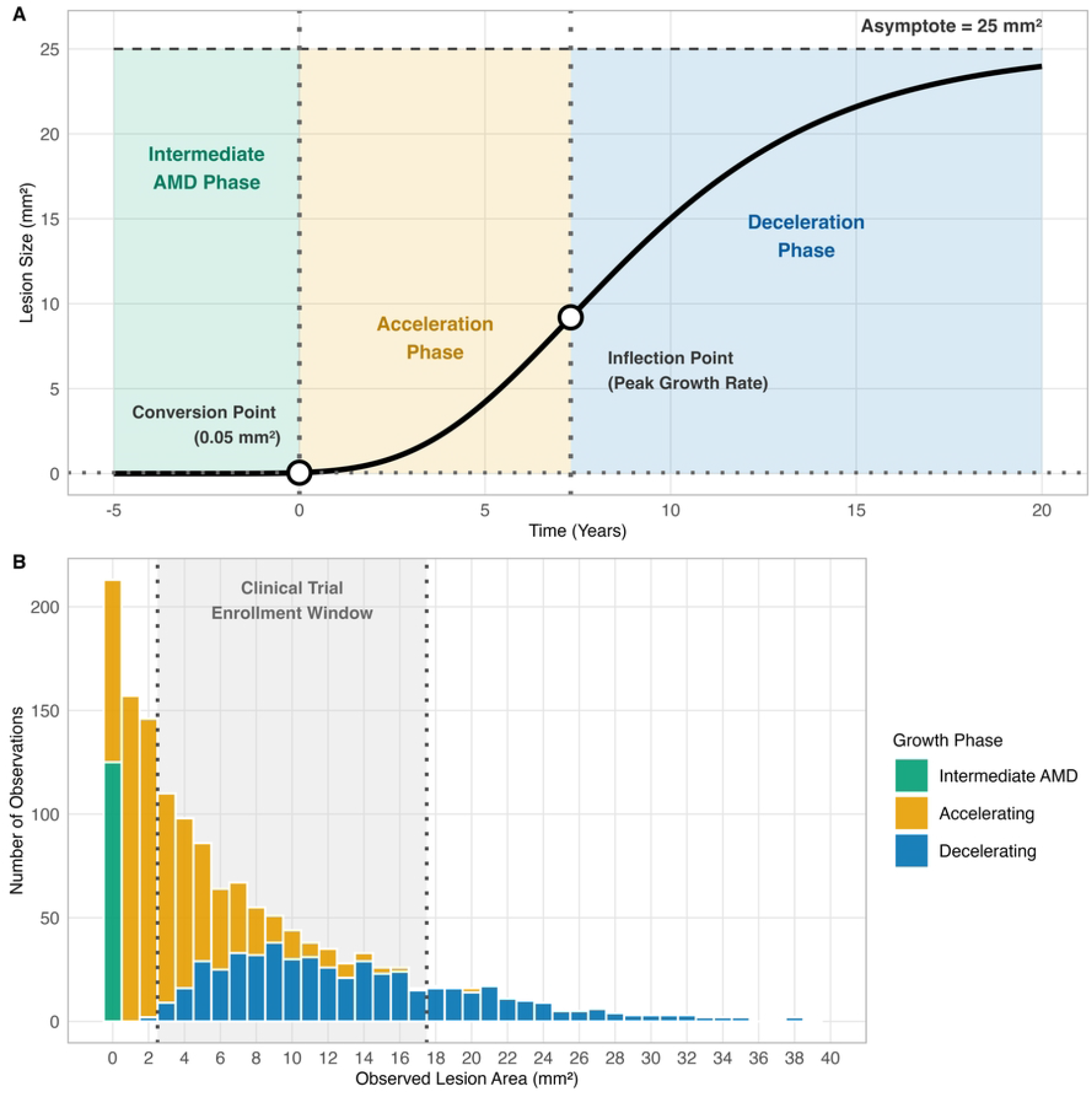
Natural growth deceleration as a potential confounder in GA RCTs. (A) Conceptual Gompertz growth curve showing acceleration to an inflection point followed by biological deceleration toward the asymptote; this inherent slowdown can mimic a treatment effect if not modeled. (B) Growth phase distribution for 1, 433 observations. The standard trial eligibility window (2.5–17.5*mm*^2^; dashed lines) spans lesions in both accelerating (amber) and decelerating (blue) phases, demonstrating heterogeneity that can bias efficacy estimates.

### Ethics approval

This retrospective study received a formal exemption from review by the Colorado Multiple Institutional Review Board (COMIRB) (Protocol #22-2198) and was conducted in accordance with the Declaration of Helsinki and applicable regulations. Because the research involved minimal risk and a secondary analysis of data collected during routine clinical care, COMIRB waived the need for patient consent. The data for this research study were accessed from July, 2023, through March, 2025. During the initial data curation process, authors had access to information that could identify individual participants, as the full dataset contained incidental protected health information. Prior to analysis, the dataset was de-identified in compliance with HIPAA, and all analyses were performed on this de-identified dataset.

## Results

### Population

Our cohort included 121 GA patients (mean age 79.2±7.9 years; 60% female), of which 49.6% had bilateral GA. At baseline, the median lesion area measured 0.8 mm^2^ (IQR, 3.8), as reported in Table 1.

**Table 1.** Summary statistics of the study cohort.

| Characteristic | Value |
| --- | --- |
| Number of Participants, n | 121 |
| Number of Eyes, n | 181 |
| Total Visits, n | 1,433 |
| Age, mean (SD), years | 79.2 (7.9) |
| Sex, n (%), F/M | 73 (60.3%) / 48 (39.7%) |
| Race, n (%) |  |
| White | 115 (95.0%) |
| Black | 1 (0.8%) |
| Asian | 1 (0.8%) |
| Other | 4 (3.3%) |
| Baseline Lesion Focality, n (%) |  |
| No GA | 49 (27.1%) |
| Unifocal | 51 (28.2%) |
| Bifocal | 29 (16.0%) |
| Multifocal (3+) | 52 (28.7%) |
| Visits per Participant, median (IQR), n | 7 (4) |
| Baseline Lesion Area, median (IQR), mm<sup>2</sup> | 0.8 (3.8) |
| Follow-up Duration, median (IQR), years | 5.8 (2.8) |

### Descriptive model performances

All models exhibited excellent convergence (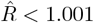; ESS *>* 1, 000; see S1-S6 Tables) [49, 50]. As illustrated in representative cases (Fig 1), the asymmetric Gompertz curve more closely tracks the observed lesion areas than the alternatives, particularly in the later-stage deceleration phase, while the logistic, linear, and effective-radius models overestimate or underestimate the data. The Gompertz model provides the best overall fit (CRPS: 0.314, SE: 0.008; MAE: 0.27), reducing CRPS by 41% relative to the linear (CRPS: 0.536, SE: 0.016; MAE: 0.52) and 35% relative to the effective-radius models (CRPS: 0.483, SE: 0.014; MAE: 0.47). Although the logistic model performs comparably in late-stage participants (CRPS: 0.321, SE: 0.009; MAE 0.28), Gompertz uniquely captures the full trajectory, including early-stage lesions. Pseudo-BMA^+^ further confirmed the superiority of the Gompertz model, assigning it a weight of 0.994 (descriptive fit) compared to negligible weights for alternative models (logistic: 0.006, linear: 0.000, effective radius: 0.000), reinforcing superior descriptive fit based on leave-one-out cross-validation (pseudo-BMA^+^ weights).

Specific parameter estimation (asymptotes, rates, and offsets) is included in S1-S6 Tables.

### Prognostic accuracy in sequential forecasting

#### Overall predictive performances

The Gompertz model outperformed all competitors across multiple forecasting metrics (S4 Fig.), with a detailed horizon breakdown in S7 Table. In rolling forecasts, the Gompertz model achieved the lowest mean CRPS (0.45 mm^2^; 95% CrI: 0.09, 1.10), followed by logistic (0.48; 0.09, 1.19), linear (0.52; 0.10, 1.26), and effective radius (0.62; 0.12, 1.49) models. Mean absolute error from predictive medians showed similar rankings: Gompertz 0.76 mm^2^ (0.68, 0.84), logistic 0.81 (0.73, 0.90), linear 0.86 (0.77, 0.96), and effective radius 1.06 (0.92, 1.20).

Using a hierarchical Gamma regression on CRPS, the posterior probability of being best was 0.923 for Gompertz (logistic 0.076; linear and effective-radius ¡0.001), indicating that, at the population level, Gompertz has the highest probability of yielding the lowest expected CRPS for a new forecast. Gompertz produced the narrowest 90% prediction intervals (2.41 mm^2^; 95% CrI 1.80–4.98) with slight under-coverage (81% vs nominal 90%; S5 Fig.). Other models were closer to nominal coverage but with wider intervals: logistic (2.73; 1.88, 6.80; 85% coverage), linear (3.99; 3.23, 6.77; 86% coverage), and effective radius (3.23; 2.71, 4.91; 87% coverage).

#### Impact of growth phase and forecast horizon on prediction

When we looked at the data by curvature phase (Table 2), we found that 61 eyes were in acceleration (33.7%), 50 eyes were in linear (27.6%), 43 eyes were in deceleration (23.8%), and 27 eyes categorized as multiple phases (14.9%). Forecasting accuracy was strongly dependent on the underlying phase, with performance differences between models most pronounced in the deceleration group. Models incapable of representing growth deceleration, specifically the effective radius and linear, performed poorly in this phase, while the Gompertz and logistic models provided the most accurate forecasts. This performance gap magnified dramatically at longer forecast horizons. In contrast, for the linear region where growth was steady, all four models performed comparably with minimal differences.

**Table 2.** Prediction accuracy varies based on a lesion’s growth pattern.

| Curvature Class | Model | Mean | 95% CrI | Prob. Best |
| --- | --- | --- | --- | --- |
| Linear Region | Linear | 0.455 | [0.366, 0.561] | 76.6% |
|  | Gompertz | 0.49 | [0.394, 0.604] | 17.5% |
|  | Effective Radius | 0.52 | [0.419, 0.640] | 3.4% |
|  | Logistic | 0.525 | [0.422, 0.646] | 2.5% |
| Acceleration | Gompertz | 0.433 | [0.335, 0.552] | 62.5% |
|  | Logistic | 0.451 | [0.349, 0.575] | 31.4% |
|  | Effective Radius | 0.488 | [0.378, 0.620] | 5.8% |
|  | Linear | 0.543 | [0.420, 0.692] | 0.2% |
| Deceleration | Gompertz | 0.429 | [0.344, 0.529] | 75.2% |
|  | Logistic | 0.454 | [0.364, 0.560] | 24.8% |
|  | Linear | 0.56 | [0.450, 0.687] | 0% |
|  | Effective Radius | 0.733 | [0.591, 0.900] | 0% |
| Multiphasic | Gompertz | 0.504 | [0.339, 0.725] | 57.8% |
|  | Logistic | 0.544 | [0.365, 0.784] | 23.3% |
|  | Linear | 0.552 | [0.372, 0.792] | 18.9% |
|  | Effective Radius | 0.819 | [0.553, 1.175] | 0% |

Prediction accuracy of the four forecasting models based on the patient’s stage of lesion growth, measured by continuous ranked probability score. The ”Prob. Best” column indicates the likelihood of each model being the most accurate. These findings highlight the Gompertz model as the most reliable tool for forecasting geographic atrophy progression in diverse clinical scenarios.

Hierarchical regression analysis revealed that forecast performance decreases with extended clinical horizons across all growth models. An additional eight months (1 SD) in forecast horizon raises forecast error by approximately 46% for Gompertz models (36%, 57%), 43% for logistic models (33%, 54%), 46% for linear models (36%, 57%), and 59% for traditional effective-radius models (48%, 71%). The Gompertz and logistic models exhibited lower degradation than the effective radius approach, showing similar resilience to longer forecast horizons.

### Impact of data sparsity

All models produced reliable forecasts even with limited historical data, demonstrating resilience to data sparsity. This low sensitivity was confirmed by hierarchical regression, which showed only a modest increase in forecast error with more observations. For each additional two visits (1 SD), the error increased by 10% for Gompertz models (2%, 18%), 9% for logistic models (1%, 17%), and 13% for both linear and effective-radius models. These findings suggest that robust forecasting does not require dense follow-up, reinforcing the clinical utility of the sigmoidal models. Although more prior visits generally improve forecasts, we observed a modest error increase of 10% (Gompertz) to 13% (linear, effective-radius) per two additional visits. This counterintuitive finding likely reflects confounding by disease severity: patients with more visits had longer follow-up and more complex disease trajectories rather than representing failure of the models to learn from additional data.

#### Case 1 (typical clinical scenario)

We validated performance on a patient selected to represent the cohort median visit count; 7 FAF images collected over 43.7 months, showing a 4.02 mm^2^ increase in GA area. When forecasting 12 months, the Gompertz model achieved an error of 0.45 mm^2^ (0.09, 1.13) versus 0.49 (0.10, 1.21) (logistic), 0.52 (0.10, 1.29) (linear), and 0.63 (0.12, 1.55) (effective radius).

#### Case 2 (sparse-data clinical scenario)

In a sparse-data scenario (4 visits, 2 years; 10 mm^2^ growth), forecasting 3 years, Gompertz achieved the lowest error of 1.22 mm^2^ (0.23, 3.05) versus 1.24 mm^2^ (0.24, 3.11) (logistic), 1.33 mm^2^ (0.26, 3.34) (linear), and 2.04 mm^2^ (0.39, 5.11) (effective radius).

### Visual validation and clinical workflow integration

This rolling validation approach reflects real-world clinical workflows, enabling physicians to update forecasts with new imaging for improved decision-making. Visual predictive checks (Fig. 3) revealed that traditional models (effective radius, linear) often overpredicted growth, especially for larger lesions and extended forecast horizons. In contrast, the Gompertz and logistic models aligned more closely with observed data. The Gompertz model maintained accuracy for large lesions (*>* 17.5 mm^2^) over 5-to-34-month forecasts, while linear models diverged significantly. Reliability improved with additional visits, as Gompertz predictions closely matched actual measurements across lesion sizes and follow-up durations.

**Fig 3.**
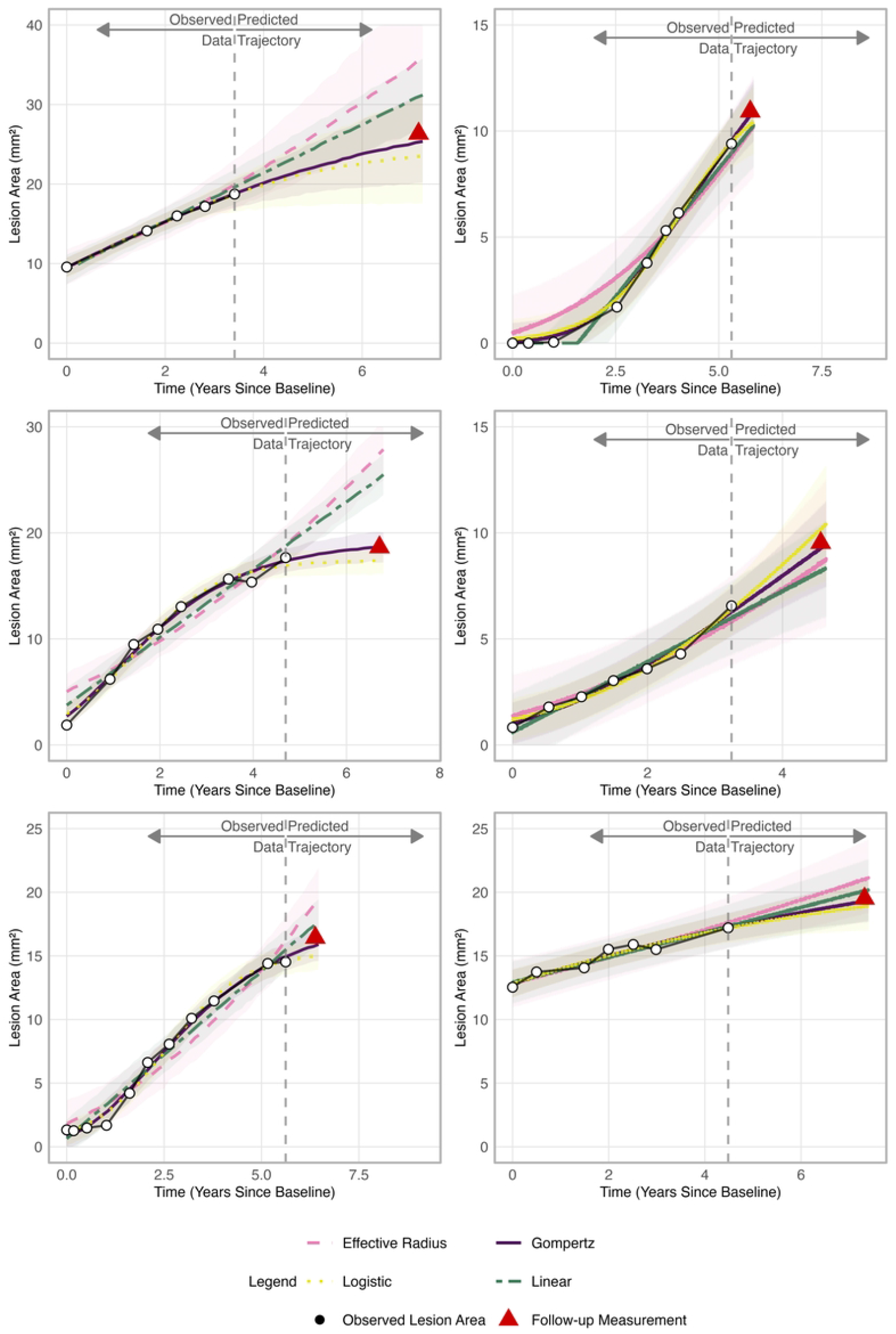
Forecast accuracy of four growth models under varying data histories and prediction horizons. One- to three-year forecasts for three eyes with varying historical visits, black circles indicate observed lesion areas (mm^2^) used for model fitting; the red triangle marks the actual follow-up measurement being predicted. Lines show posterior means and shaded 90% credible intervals for effective radius (pink), Gompertz (purple), linear (green), and logistic (yellow) models. Across all six scenarios, the Gompertz model produces predictions closest to the observed value, whereas effective-radius and linear models systematically overpredict, an effect most pronounced at longer horizons. Logistic performance is intermediate.

## Discussion

In our cohort of 121 GA patients, the Gompertz model provided the most accurate and reliable forecasts, outperforming all competitors by achieving the lowest forecast error (CRPS: 0.45 mm^2^). While the absolute difference in mean CRPS is modest (0.45 vs 0.62 mm^2^), the clinical impact of this 27% relative improvement becomes more significant over longer, multi-year forecast horizons where the absolute errors of traditional models diverge more sharply. Furthermore, the Gompertz model maintained superior predictive stability over time, with a lower rate of error accumulation (32% per 8 months) than traditional effective-radius models (59%), allowing for more reliable long-term predictions.

Similar sigmoidal models have been well-established in other specialties [25, 39], yet adoption in GA research has lagged. Gompertz model superiority stems from its biological realism; its asymmetric sigmoidal shape better captures GA’s natural history, an initial phase of rapid enlargement followed by deceleration as the lesion approaches anatomical boundaries. In contrast, unbounded models such as the linear and square root systematically overpredict lesion expansion, which can inflate expected control-arm progression and dilute apparent treatment effects. Our analysis confirms recent evidence that the widely used square-root transformation fails to eliminate baseline-size dependence and performs worse than even a simple linear fit [29]. As the first robust application of Gompertz modeling to longitudinal GA data, this study fills a critical methodological gap and adds a novel dimension to understanding GA lesion progression (S3 Fig.).

### RCT implications

Our analysis reveals that a patient’s natural growth phase is not merely random error, but a powerful, unadjusted confounder with a systematic distribution in the trial population itself. This is particularly critical for current trials of complement inhibitors, such as the OAKS/DERBY (Syfovre, pegcetacoplan) [2] and GATHER trials (Izervay, avacincaptad pegol) [3, 4], which enroll patients with lesions up to 17.5 mm^2^. As visualized in Fig. 2B, we found that an unbalanced proportion of trial-eligible visits (42%) were already in a post-inflection, deceleration phase. For patients near this upper limit, any observed slowdown is ambiguous, attributable to either treatment or the lesion’s natural life cycle. This pattern could bias interpretation by conflating natural deceleration with treatment effects, confounding trial outcomes.

By leveraging Gompertz modeling, trial sponsors could address this directly by tailoring recruitment based on patient-specific growth trajectories, ensuring participants are enrolled during disease phases where intervention impact is most measurable. The model’s capability to predict biological asymptotes also provides a useful benchmark for interpreting control-arm progression, potentially reducing sample size requirements while increasing statistical power to detect meaningful treatment differences. This approach may increase trial sensitivity, accelerate approvals and improve resource efficiency.

Furthermore, the Gompertz model’s parameters, the asymptotic area (biological limit) and the deceleration rate, reveal significant inter-patient and inter-eye variability. This paves the way for personalized prognoses and adaptive trial designs targeting specific subgroups most likely to benefit from treatment. Accurately identifying the growth inflection point could also optimize the timing of therapeutic interventions, which may be most effective before the lesion enters its plateau phase.

### Clinical applications

A key clinical strength of the Gompertz model is its resilience to the data sparsity common in real-world settings. Our forecasting validation demonstrates that it robustly predicts lesion trajectories even with irregular follow-up or incomplete imaging histories. This resilience stems from the model’s strong biological foundation; by assuming a plausible sigmoidal trajectory, it can generate reliable long-term forecasts where simpler models cannot. The hierarchical Bayesian framework further enhances its adaptability by accounting for patient-specific variability in disease progression, resulting in predictions that are clinically actionable.

Integrating Gompertz predictions into patient conversations can also improve shared decision-making. Personalized timelines based on the model’s outputs help patients better understand their disease progression and evaluate treatment options, including participation in RCTs. This patient-centered approach aligns with precision medicine principles and demonstrates how mathematical modeling can enhance routine ophthalmic care through clearer communication of prognosis and treatment timing.

### Limitations and future directions

Automated FAF segmentation may introduce systematic measurement bias [34], particularly in areas of low signal where lesion boundaries are ambiguous, despite our quality control procedures. The six-month imaging interval limits detection of short-term growth inflections, while our reliance on FAF rather than more clinically accessible OCT-derived GA measurements may reduce translational relevance. Our single-center, predominantly Caucasian cohort may not generalize to diverse populations. Finally, while effective for modeling bounded growth trajectories, the Gompertz model cannot capture complex growth phenomena including spatial heterogeneity, satellite lesion formation, or discontinuous expansion patterns.

Future work should extend the Gompertz framework by integrating spatially resolved photoreceptor metrics with biologic covariates, such as complement activation profiles and GA-associated risk variants [51–53], to further refine prognostication, uncover modifiable disease mechanisms, and enable patient stratification. Coupling these biologically informed parameters with spatial predictions of lesion proximity to the fovea will bridge the gap between lesion-area metrics and functional outcomes (e.g., visual-acuity loss) that recent studies have highlighted [54–56], thereby guiding risk-based therapeutic interventions.

The integration of Gompertz modeling into routine clinical workflows and trial frameworks represents a key translational goal. Practical applications include real-time forecasting to improve treatment timing and reconstructing historical growth patterns to refine estimates of disease milestones. These predictive tools could ultimately set the stage for novel strategies in GA management, from preventative interventions to retrospective natural history studies.

In conclusion, the common use of unbounded models overestimates GA progression and can mislead trial interpretation. By reflecting the natural deceleration of lesion growth, the Gompertz curve provides superior prognostic accuracy in CORIS. Integrating this biologically grounded modeling into clinical workflows and trial design is a key translational goal, representing a significant advancement that enables more efficient therapeutic evaluations and sets more realistic, personalized expectations for disease progression.

## Data Availability

The clinical data underlying this study contain protected health information collected during routine care and cannot be shared publicly under HIPAA and institutional policy. De-identified data may be requested from the Department of Ophthalmology’s research administration office at. Requests must include a brief research proposal and will require IRB approval and/or a Data Use Agreement as determined by institutional compliance. Access decisions are made by institutional offices independent of the authors; the study authors do not make access determinations but may provide technical clarification upon request.

All analysis code, model specifications, and variable definitions are publicly available at https://github.com/CodeByAaron/Geographic-Atrophy-Lesion-Growth.

## Funding/Support

This project was supported in part by an Unrestricted Research Grant to the Department of Ophthalmology from Research to Prevent Blindness and by the NIH/NCATS Colorado CTSA (Grant UM1 TR004399). The funding organizations had no role in the design and conduct of the study, the collection, management, analysis, and interpretation of the data, the preparation, review, or approval of the manuscript, or the decision to submit the manuscript for publication.

## Competing interests

Dr. McNamara reports consulting for Evolution Optiks. Dr. Manoharan reports clinical-trial support from Iveric Bio and Regeneron and consulting for Genentech. Dr. Mandava reports consulting for Soma Logic and ONL Therapeutics and holds a patent with Alcon. Dr. Kalpathy–Cramer reports equity in Siloam Vision, grant support from GE Healthcare and Genentech, and a pending patent for a retinopathy-of-prematurity deep-learning algorithm. Dr. de Carlo Forest reports contractor services for Genentech. No other competing interests were declared by the remaining authors (A. D. Beckwith, Y. A. Veturi, B. Bearce, S. Kinder, R. Gnanaraj, A. Lynch, P. Singh, G. Nebbia).

## Author Contributions

**Conceptualization:** Beckwith, Kalpathy-Cramer, Mandava, Nebbia, and McNamara. **Data curation:** Beckwith, Veturi, Manoharan, de Carlo Forest, Gnanaraj, Kinder, Bearce. **Formal analysis:** Beckwith, Kalpathy-Cramer. **Investigation:** Manoharan, de Carlo Forest, Gnanaraj, Lynch. **Methodology:** Beckwith, Kalpathy-Cramer, Nebbia, and McNamara. **Project administration:** Bearce, Mandava, Kalpathy-Cramer. **Resources:** Mandava, Kinder, Bearce, Kalpathy-Cramer. **Software:** Veturi, Kinder, Bearce. **Supervision:** Mandava, Kalpathy-Cramer, Singh, Nebbia, McNamara. **Validation:** Beckwith, Nebbia, McNamara, Veturi, Kalpathy-Cramer. **Visualization:** Beckwith, Veturi. **Writing – original draft:** Beckwith, McNamara, Veturi. **Writing – review and editing:** All authors.

Beckwith and Veturi had full access to all the data in the study and take responsibility for the integrity and accuracy of the data analysis.

## Acknowledgments

Rolling forecast computations were performed on the Alpine high performance computing cluster at the University of Colorado Boulder, jointly funded by the University of Colorado Boulder, the University of Colorado Anschutz, and Colorado State University with support from NSF Grants OAC 2201538 and OAC 2322260.

## Supporting information

**S1 Fig. Cohort assembly and data flow**.This flowchart illustrates the cohort selection process. A key quality control step involved flagging potential outliers using isotonic regression residuals; this method was chosen to avoid biasing the final cohort toward any particular growth model. Because isotonic regression only assumes that lesion growth is monotonic (i.e., does not shrink over time), it can identify biologically implausible data points indicative of segmentation errors without making assumptions about whether the true growth pattern is linear or sigmoidal. All cases flagged by this method were subsequently visually verified by our team.

**S2 Fig. Bounded vs. unbounded GA area growth under a common parameterization (asymptote** = 25**; growth rate** *≈* 0.4).Six mathematical growth models (Gompertz, logistic, Bertalanffy, Mitscherlich, effective radius, and linear), each parameterized to approach a similar final lesion size (asymptote = 25). The x-axis is shifted to highlight how each model’s assumptions lead to distinct early- and late-phase growth behaviors, from unbounded linear expansion to varying forms of sigmoidal saturation.

**S3 Fig. Geographic atrophy growth trajectories reveal underlying common shape through progressive alignment**.Panel A shows raw lesion area trajectories (mm^2^) over time, illustrating substantial heterogeneity in baseline size, growth rate, and follow-up duration. Panel B displays the same trajectories after L^2^ normalization and optimal time alignment, revealing a conserved sigmoidal growth pattern across the cohort. Panel C presents trajectories aligned using first-jerk registration (third derivative = 0), which identifies a biologically meaningful inflection point that can serve as a prognostic landmark for clinical timing. The progression from apparent chaos (A) to revealed order (B) to clinical utility (C) demonstrates that patient-to-patient variability largely reflects differences in disease timing rather than fundamentally different growth processes.

**S4 Fig. Posterior evidence: pairwise differences in continuous ranked probability scores**, Δ**CRPS (mm**^2^**), for rolling forecasts, ridgeline densities comparing models**. We arranged ΔCRPS such that lower CRPS reflects better probabilistic predictions (combining accuracy and interval calibration), favoring the model listed first. Vertical markers denote posterior medians; shaded bands highlight central credible intervals. The leftward shift and mass below zero indicate consistently better calibrated and more accurate forecasts under the Gompertz model.

**S5 Fig. Adjusted probability-integral-transform (PIT) histograms for four candidate growth models**. Histograms display the distribution of PIT values after decorrelating within-eye predictions, a perfectly calibrated model would yield a flat histogram at the reference frequency (red dashed line). Gompertz and Logistic curves produce nearly uniform PITs with only a mild central mound indicating slight under-dispersion, whereas the Linear model shows a pronounced spike at 0.5 and the effective radius model is left-skewed (systematic over-prediction). Overall, Gompertz *≈* Logistic *≫* Effective-Radius *>* Linear for calibration quality.

**S1 Table. Gompertz model posterior distribution statistics**. Parameter estimates, standard errors, 95% credible intervals, and effective sample sizes for the hierarchical Gompertz growth model. Population-level parameters (*A*_pop_, *b*_pop_, *c*_pop_) represent asymptotic lesion size, offset parameter, and growth rate respectively. Random effects capture patient-level and patient-laterality-level variation with correlation parameters showing relationships between growth parameters. All chains showed adequate convergence 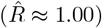 with sufficient effective sample sizes.

**S2 Table. von Bertalanffy model posterior distribution statistics**. Complete posterior summary for the hierarchical von Bertalanffy growth model with identical structure to the Gompertz parameterization. The model demonstrates good convergence properties and captures similar hierarchical variation patterns as the Gompertz model, though with different population-level parameter estimates reflecting the distinct mathematical form of the von Bertalanffy curve.

**S3 Table. Logistic model posterior distribution statistics**. Posterior estimates for the hierarchical logistic growth model showing population-level parameters and random effect structures. The logistic model’s symmetric S-curve produces different parameter estimates compared to the asymmetric Gompertz and von Bertalanffy models, particularly in the growth rate and offset parameters, while maintaining similar hierarchical variance components.

**S4 Table. Mitscherlich model posterior distribution statistics**. Posterior summary for the hierarchical Mitscherlich (monomolecular) growth model. This model shows the poorest fit among the sigmoidal models, reflected in larger residual variance and different parameter scaling. The Mitscherlich form assumes initially linear growth transitioning to asymptotic behavior, which appears less compatible with observed GA progression patterns.

**S5 Table. Linear model posterior distribution statistics**. Posterior estimates for the hierarchical linear growth model serving as the baseline comparison. Parameters represent intercept (*β*_0_) and slope (*β*_time_) with patient-level and patient-laterality-level random effects. The linear model’s constant growth assumption results in substantially larger residual variance compared to sigmoidal alternatives, indicating poorer fit to the deceleration observed in GA progression.

**S6 Table. Effective radius model posterior distribution statistics**. Posterior summary for the hierarchical effective radius model assuming circular lesion geometry with linear radius expansion. Parameters *b*_0_ and *b*_1_ represent baseline radius and radius growth rate respectively. This geometric constraint produces the poorest overall fit, with the highest residual variance reflecting the mismatch between assumed circular growth and actual irregular GA lesion morphology.

**S7 Table. Comprehensive forecasting performance metrics across prediction horizons**. Detailed comparison of predictive accuracy using continuous ranked probability scores (CRPS) and mean absolute error (MAE) at multiple forecast horizons from 4 months to 3 years. The Gompertz model consistently demonstrates superior forecasting performance across all time horizons, with the CRPS-based posterior probability that Gompertz is best equal to 0.923. Predictive factors quantify how forecast accuracy degrades with longer prediction horizons and sparser data availability.

**S8 Table. Additional model comparison metrics for Bertalanffy and Mitscherlich models**. Extended leave-one-out cross-validation results showing expected log pointwise predictive density (ELPD), effective number of parameters (*P*_loo_), mean absolute error (MAE), and pseudo-Bayesian model averaging weights. These metrics confirm the Gompertz model’s superiority (pseudo-BMA^+^ weight = 0.994) while showing that Bertalanffy performs second-best among the sigmoidal alternatives, with Mitscherlich showing substantially poorer fit to the data.

**S1 Appendix. Cohort characteristics & Bayesian balance checks**

**S2 Appendix. Mathematical formulations of comparator growth models S3 Appendix. Prior specification, sampler settings, & diagnostics**

## Notes

### Competing Interest Statement

I have read the journal's policy and the authors of this manuscript have the following competing interests: Dr. McNamara reports consulting for Evolution Optiks. Dr. Manoharan reports clinical-trial support from Iveric Bio and Regeneron and consulting for Genentech. Dr. Mandava reports consulting for Soma Logic and ONL Therapeutics and holds a patent with Alcon. Dr. Kalpathy--Cramer reports equity in Siloam Vision, grant support from GE Healthcare and Genentech, and a pending patent for a retinopathy-of-prematurity deep-learning algorithm. Dr. de Carlo Forest reports contractor services for Genentech. No other competing interests were declared by the remaining authors (A. D. Beckwith, Y. A. Veturi, B. Bearce, S. Kinder, R. Gnanaraj, A. Lynch, P. Singh, G. Nebbia).

### Author Declarations

This retrospective study was approved by the Colorado Multiple Institutional Review Board.

